# Genetic predisposition for developmental dyslexia in semantic variant primary progressive aphasia

**DOI:** 10.1101/2025.10.10.25337733

**Authors:** Sterre C.M. de Boer, Jenna Najar, Jochum J. van ’t Hooft, Chenyang C. Jiang, Kim M.A. van Rooijen, Per Östberg, Sven J. van der Lee, Betty Tijms, Colin Groot, Afina W. Lemstra, Yolande A.L. Pijnenburg, Lianne M. Reus, Alexander F. Santillo

## Abstract

**Importance:** Mechanisms behind the asymmetric left-hemispheric atrophy observed in primary progressive aphasia (PPA), a neurodegenerative disorder related to either frontotemporal lobar degeneration or Alzheimer’s disease, remains a mystery. There is an increased prevalence of dyslexia in PPA patients and their relatives, especially in the logopenic variant PPA (lvPPA). This co-occurrence questions whether genetic variants associated with dyslexia are linked to selective neurodegeneration in language-associated brain regions.

**Objective:** To understand if genetic risk for dyslexia contributes to selective vulnerability in PPA, we determined whether polygenic risk scores (PRS) for dyslexia are elevated in PPA in comparison to other dementias and healthy controls. Given previous literature, we hypothesized that PRS for dyslexia is highest in individuals with lvPPA.

**Participants:** Genotyped participants from Amsterdam Dementia Cohort (ADC) diagnosed with semantic variant PPA (svPPA), non-fluent variant PPA (nfvPPA), lvPPA, right-temporal variant FTD (rtvFTD), PPA not otherwise specified (PPA NOS), behavioral variant FTD (bvFTD), Alzheimer’s dementia (AD) and dementia with Lewy bodies (DLB), in addition to controls, were included. Sex, age at blood sampling and education was collected. For each participant, PRS for dyslexia were created using genome-wide association study summary statistics on dyslexia (n = 51,800 cases and 1,087,070 controls, a study from 23andMe, Research Institute.). PRS were computed with LDpred2 and standardized.

**Main outcome(s) and Measure(s):** Differences in PRS across diagnostic groups were assessed using linear regression models, with age, sex, education, diagnosis, and the diagnosis × sex interaction term as predictors. Since interaction term diagnosis*sex was significant, analysis was stratified for sex.

**Results:** A total of 3,407 ADC participants were included, of which n=969 controls, n=66 svPPA, n=30 nfvPPA, n=38 lvPPA, n=51 rtvFTD, n=17 PPA NOS, n=230 bvFTD, n=1750 AD and n=256 DLB cases. Within males, PRS for dyslexia were significantly higher in svPPA compared to all other diagnoses and controls (all p-values <0.05), except compared to nfvPPA. Within females, no significant differences emerged.

**Conclusion and Relevance:** Our findings reveal a shared genetic basis between neurodevelopmental dyslexia and svPPA, indicating a link between developmental and neurodegenerative language disorders.

## Introduction

The pronounced left-hemispheric brain atrophy observed in the neurodegenerative disorder primary progressive aphasia (PPA), a clinical syndrome characterized by a gradual increase of language difficulties, demonstrates that neurodegenerative processes can selectively target language-dominant brain regions [1]. Yet, the reasons for vulnerability of these brain regions to neurodegenerative processes remains poorly understood.

PPA is currently classified into three major subtypes: the logopenic variant PPA (lvPPA), which is most often associated with Alzheimer’s disease (AD) pathology, and the nonfluent and semantic variants of PPA (nfvPPA, svPPA respectively), both of which are linked to frontotemporal lobar degeneration pathology (FTLD) [1]. Notably, individuals with PPA show a disproportionately high prevalence of neurodevelopmental learning disorders in their medical and family history, particularly developmental dyslexia [2]. A genetic link between dyslexia and AD has been described [3] and among PPA subtypes, dyslexia prevalence was highest in lvPPA [4]. From a clinical perspective, lvPPA and developmental dyslexia also overlap in disease characteristics such as phonological processing difficulties [1, 5]. Anatomically, a pathological case series of lvPPA patients demonstrated that neurodevelopmental abnormalities and AD pathology also overlapped within perisylvian regions, areas implicated in both dyslexia and lvPPA [4, 6].

Given prior evidence suggesting a link between neurodevelopmental disorder dyslexia and neurodegeneration later in life, we theorize that genetic predisposition for dyslexia might contribute to the selective temporal lobe vulnerability observed in PPA. Polygenic risk scores (PRS), which combine the cumulative effects of common genetic variance associated with a trait, provide a tool to assess this proposed genetic liability for dyslexia across neurodegenerative disorders. Based on prior evidence, we hypothesized that PRS for dyslexia is increased in PPA, particularly lvPPA.

## Methods

A total of 3,407 individuals were selected from the Amsterdam Dementia Cohort (ADC). A detailed cohort description can be found in the supplement. In short, the study sample consisted of patients with FTD-related syndromes including semantic variant PPA (svPPA), non-fluent variant (nfvPPA), right temporal variant FTD (rtvFTD) and behavioral variant FTD (bvFTD) as well as those with AD-related syndromes including lvPPA and AD-type dementia. Other clinical subtypes included were dementia with Lewy bodies (DLB) and PPA not otherwise specified (PPA NOS) in addition to controls. Clinical data collection comprised sex, age at blood sampling and education (in years). Genotyping was performed using Illumina Global Screening Array (GSA), and imputation with TOPMed, as described previously [7].

PRS for dyslexia were created using genome-wide association study (GWAS) summary statistics on dyslexia (n = 51,800 cases and 1,087,070 controls, a study from 23andMe Research Institute, a consumer genetics company) [8]. PRS were computed with LDpred2, a Bayesian approach that captures the polygenic nature of traits, such as dyslexia [9]. All PRS were standardized (mean=0, SD=1) (see supplements for more methodological information). Differences in PRS (outcome) across diagnostic groups were assessed using linear regression models, with age, sex, education, diagnosis, and the diagnosis × sex interaction term as predictors. Results were reported separately for males and females, as the interaction term diagnosis × sex was significant. All analysis were performed using R (version 4.3.1).

## Results

Demographic characteristics are shown in Table 1. Within males only, PRS for dyslexia were significantly higher in svPPA patients compared to all other diagnoses and controls (all p-values <0.05), except compared to the nfvPPA group (Table 2, Figure 1). Within females, no significant differences emerged. Results without sex stratification were similar to those in males, with significant higher PRS for dyslexia in the svPPA group compared to all other diagnostic groups, except for lvPPA (Supplementary Material Table S1).

**Table 1.** Demographics.

|  | *Total sample*<br>*N=3,407* | **Controls**<br>**N=969** | **svPPA**<br>**N=66** | **nfvPPA**<br>**N=30** | **lvPPA**<br>**N=38** | **rtvFTD**<br>**N=51** | **PPA NOS**<br>**N=17** | **bvFTD**<br>**N=230** | **AD**<br>**N=1750** | **DLB**<br>**N=256** | **p-value** |
| --- | --- | --- | --- | --- | --- | --- | --- | --- | --- | --- | --- |
| **Age years,**<br>**mean (SD)** | *63.8 (8.6)* | 59.2 (8.5) | 64.1 (5.7) | 67.9 (7.9) | 68.8 (7.0) | 63.3 (6.5) | 67.9 (5.8) | 62.7 (9.0) | 65.5 (7.9) | 68.8 (6.8) | 1.85e-101<sup>a</sup> |
| **Female, N**<br>**(%)** | *1,514*<br>*(44.4%)* | 377 (38.9%) | 24 (36.4%) | 17 (56.7%) | 19 (50.0%) | 14 (27.5%) | 8 (47.1%) | 93 (40.4%) | 917 (52.4%) | 45 (17.6%) | 4.93e-27<sup>b</sup> |
| **Education**<br>**years\*,**<br>**median**<br>**[IQR]** | *13.0 [11.0-*<br>*15.0]* | 13.0 [13.0-15.0] | 13.0 [11.0-15.0] | 11.0 [11.0-13.0] | 13.0 [11.0-15.0] | 13.0 [11.0-15.0] | 13.0 [13.0-15.0] | 13.0 [11.0-14.0] | 13.0 [11.0-15.0] | 13.0 [11.0-15.0] | 4.77e-12<sup>c</sup> |

Abbreviations: AD, Alzheimer’s disease; bvFTD, behavioral variant frontotemporal dementia; DLB, dementia with Lewy bodies; nfvPPA, non-fluent variant primary progressive aphasia; svPPA, semantic variant primary progressive aphasia; lvPPA, logopenic variant primary progressive aphasia; PPA NOS, primary progressive aphasia not otherwise specified; SD, standard deviation; IQR, interquartile range. \* 84 missing values.
<sup>a</sup>One-way ANOVA
<sup>b</sup>Chi-square
<sup>c</sup>Kruskall-Wallis

**Table 2.** Linear regression analysis stratified for sex.

| Sex | Reference Group | Diagnostic Group | Estimate | Standard Error | p-value |
| --- | --- | --- | --- | --- | --- |
| M | svPPA | rtvFTD | -0.401 | 0.118 | 6.84e-04* |
|  |  | lvPPA | -0.293 | 0.144 | 4.20e-02* |
|  |  | nfvPPA | -0.291 | 0.165 | 7.82e-02 |
|  |  | PPA NOS | -0.551 | 0.191 | 3.88e-03* |
|  |  | controls | -0.373 | 0.085 | 1.10e-05* |
|  |  | bvFTD | -0.330 | 0.093 | 3.99e-04* |
|  |  | AD | -0.376 | 0.084 | 7.46e-06* |
|  |  | DLB | -0.383 | 0.09 | 2.02e-05* |
| F | svPPA | rtvFTD | -0.015 | 0.178 | 9.33e-01 |
|  |  | lvPPA | 0.024 | 0.158 | 8.80e-01 |
|  |  | nfvPPA | -0.067 | 0.170 | 6.95e-01 |
|  |  | PPA NOS | -0.329 | 0.210 | 1.18e-01 |
|  |  | controls | -0.111 | 0.109 | 3.11e-01 |
|  |  | bvFTD | 0.066 | 0.119 | 5.79e-01 |
|  |  | AD | 0.023 | 0.107 | 8.26e-01 |
|  |  | DLB | 0.079 | 0.132 | 5.51e-01 |
| M | nfvPPA | rtvFTD | -0.11 | 0.167 | 5.08e-01 |
|  |  | lvPPA | -0.003 | 0.186 | 9.87e-01 |
|  |  | PPA NOS | -0.261 | 0.224 | 2.44e-01 |
|  |  | controls | -0.083 | 0.145 | 5.69e-01 |
|  |  | bvFTD | -0.039 | 0.150 | 7.93e-01 |
|  |  | AD | -0.085 | 0.144 | 5.54e-01 |
|  |  | DLB | -0.092 | 0.148 | 5.33e-01 |
| F | nfvPPA | rtvFTD | 0.051 | 0.196 | 7.93e-01 |
|  |  | lvPPA | 0.090 | 0.178 | 6.12e-01 |
|  |  | PPA NOS | -0.263 | 0.226 | 2.45e-01 |
|  |  | controls | -0.044 | 0.138 | 7.48e-01 |
|  |  | bvFTD | 0.132 | 0.145 | 3.61e-01 |
|  |  | AD | 0.090 | 0.135 | 5.04e-01 |
|  |  | DLB | 0.145 | 0.155 | 3.49e-01 |
| M | lvPPA | rtvFTD | -0.107 | 0.146 | 4.62e-01 |
|  |  | PPA NOS | -0.258 | 0.209 | 2.17e-01 |
|  |  | controls | -0.080 | 0.121 | 5.12e-01 |
|  |  | bvFTD | -0.036 | 0.127 | 7.74e-01 |
|  |  | AD | -0.083 | 0.120 | 4.92e-01 |
|  |  | DLB | -0.089 | 0.124 | 4.71e-01 |
| F | lvPPA | rtvFTD | -0.039 | 0.186 | 8.35e-01 |
|  |  | PPA NOS | -0.353 | 0.217 | 1.04e-01 |
|  |  | controls | -0.135 | 0.122 | 2.72e-01 |
|  |  | bvFTD | 0.042 | 0.131 | 7.48e-01 |
|  |  | AD | 0 | 0.119 | 9.98e-01 |
|  |  | DLB | 0.055 | 0.142 | 6.99e-01 |
| M | rtvFTD | PPA NOS | -0.151 | 0.192 | 4.33e-01 |
|  |  | controls | 0.028 | 0.088 | 7.53e-01 |
|  |  | bvFTD | 0.071 | 0.096 | 4.59e-01 |
|  |  | AD | 0.025 | 0.087 | 7.75e-01 |
|  |  | DLB | 0.018 | 0.092 | 8.45e-01 |
| F | rtvFTD | PPA NOS | -0.314 | 0.232 | 1.75e-01 |
|  |  | controls | -0.096 | 0.145 | 5.10e-01 |
|  |  | bvFTD | 0.081 | 0.153 | 5.97e-01 |
|  |  | AD | 0.039 | 0.144 | 7.89e-01 |
|  |  | DLB | 0.094 | 0.164 | 5.68e-01 |

Adjusted for age and education (years). \* statistically significant.

**Figure 1.**
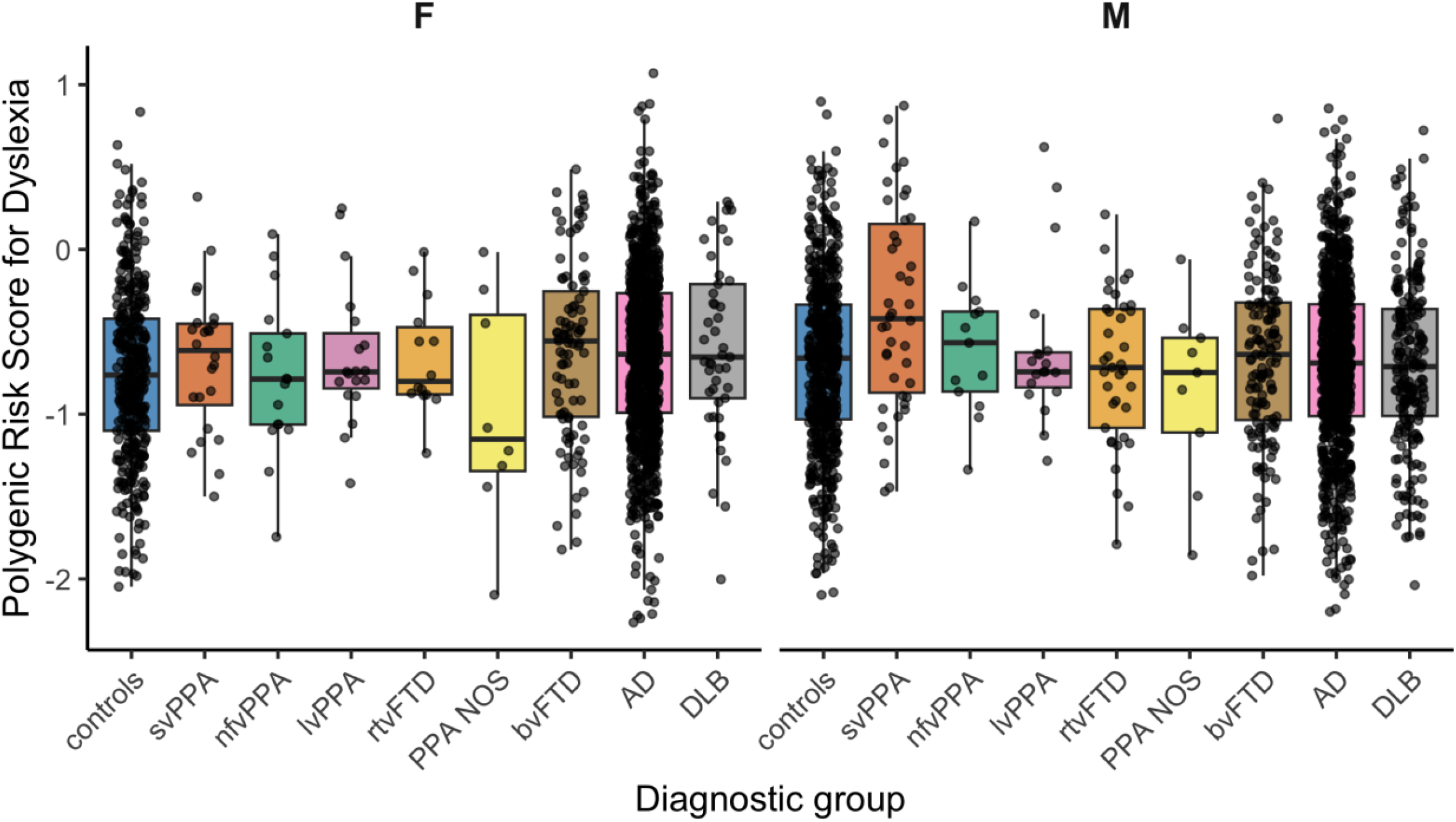
Polygenetic score for dyslexia by diagnostic group, stratified by sex. In the male group (right), PRS for dyslexia is significantly higher in svPPA compared to all other groups, except to nfvPPA. In the female group (left), no significant differences were found. Abbreviations: AD, Alzheimer’s disease; bvFTD, behavioral variant frontotemporal dementia; DLB, dementia with Lewy bodies; F, female; nfvPPA, non-fluent variant primary progressive aphasia; svPPA, semantic variant primary progressive aphasia; lvPPA, logopenic variant primary progressive aphasia; M, male; PPA NOS, primary progressive aphasia not otherwise specified.

## Discussion

In contrast to our hypothesis that dyslexia PRS would be elevated in lvPPA, we found increased PRS for developmental dyslexia in svPPA which suggests that the effects of a genetic predisposition for developmental dyslexia extend across the entire lifespan, with its neurobiological imprint affecting both neurodevelopment and frontotemporal lobar degeneration.

The reason why dyslexia PRS are increased specifically in svPPA is intriguing. Whereas lvPPA is characterized by phonological processing deficits, a core characteristic also seen in dyslexia, svPPA is characterized by anomia reflecting a transmodal semantic deficit and a pattern of surface dyslexia, in which phonological decoding is intact [1, 10]. Current dyslexia models implicate both phonological and orthographic decoding deficits, the latter especially relevant in nontransparent (e.g., English) or logographic (e.g., Chinese) writing systems [10, 11]. Our findings suggest these decoding processes rely more on semantic mechanisms than previously thought, offering a novel contribution to the dyslexia-model debate. From a pathological perspective, svPPA is characterized by predominantly left-sided anterior temporal lobe atrophy, which is often disproportionately severe compared with the clinical profile observed, and its strong clinico-pathological association with TAR DNA-binding protein 43 type C (TDP-43) [12, 13]. Interestingly, dyslexia susceptibility genes have been associated with cortical thickness reduction in language regions in patients with FTD [14]. Neural correlates of dyslexia have been localized to the temporoparietal and perisylvian regions, as well as to occipitotemporal networks, often referred to as the ventral or “what” stream of perception, which are strongly implicated in svPPA [6]. It is therefore plausible that genetic predisposition for dyslexia leads to early vulnerability of the temporal lobe and its network, priming neuronal cell loss and rendering it structurally more vulnerable and thereby predisposing it to be the first region affected in svPPA.

The observed sex difference necessitates further investigation and discussion. The original GWAS included sex-stratified analyses and the results were used to calculate sex-specific PRS for dyslexia which did not alter the results (Supplementary Material S2). Although literature suggests that dyslexia is more prevalent in males [15], the original GWAS reported comparable prevalences between females (4.5%) and males (4.6%) [8]. One possible explanation is that males might be more thoroughly assessed for dyslexia, resulting in more accurate self-reported diagnoses in 23andMe.

Secondly, the original GWAS study identified one genome-wide significant variant (*TMEM25*) on the X chromosome. Sex chromosomes were excluded from our PRS calculation. Females (who carry two X chromosomes) may therefore, in theory, have lower calculated PRS in our study. Alternatively, this pattern may reflect a genuine, yet elusive, sex-specific genetic link between dyslexia and svPPA present only in males.

This study is not without limitations. First, 23andMe’s research-consented cohort is susceptible to selection bias, with self-reported dyslexia diagnoses that may involve misreporting or unknown proneness to forms of dyslexia. Second, sample size of each dementia subtype varied and were, in some groups, small. Replication of our findings is therefore warranted. Third, the prevalence of dyslexia in our patient and control groups were unknown. The closest proxy for dyslexia available was education. We conducted sensitivity analyses without adjustment for education, showing similar results of higher dyslexia PRS for svPPA compared to all other groups except lvPPA and nfvPPA (Supplementary Material S3). It is possible that a higher proportion of patients with svPPA had dyslexia, which could account for the elevated PRS. This in itself is noteworthy, as it would provide genetic evidence supporting a higher prevalence of dyslexia among patients with svPPA.

## Conclusion

Taken altogether, we provide the first evidence that polygenic risk for dyslexia is increased in a neurodegenerative language disorder of adulthood, namely svPPA. Our results frame neurodevelopmental language disorders into a lifespan perspective, offering a novel clue into the developmental underpinnings of svPPA specifically and neurodegenerative conditions in general.

## Data Availability

All data produced in the present study are available upon reasonable request to the authors.

## Acknowledgments

Participating patients and families of Amsterdam Dementia Cohort (ADC), staff and colleagues at Alzheimercentrum Amsterdam. Additionally, we would like to thank the research participants and employees of 23andMe Research Institute for making this work possible.

