## supplements for "Genetic predisposition for developmental dyslexia in semantic variant primary progressive aphasia"

### Genetic predisposition for developmental dyslexia in semantic variant primary progressive aphasia - Online supplementary material

#### S1. Supplementary cohort description and ethics

Participants from the Amsterdam Dementia Cohrt

The Amsterdam Dementia Cohort (ADC) is an ongoing longitudinal, observational study initiated in 2000 at the Alzheimer Center Amsterdam, a tertiary dementia clinic part of the AmsterdamUMC, location VUmc (prior called VU University Medical Center) [1, 2]. The cohort comprises individuals referred for diagnostic evaluation of cognitive complaints, encompassing a broad spectrum of both neurodegenerative and non-neurodegenerative disorders. All patients referred to the Alzheimer Center for diagnostic assessment are eligible for inclusion in the ADC. Diagnostic work-up include extensive history taking, neurological examination, neuropsychological assessment, structural brain MRI, electrophysiological recordings and determination of Alzheimer's Disease biomarkers in cerebrospinal fluid (CSF) or Amyloid PET-scans. Blood (serum and plasma) and DNA sampling are biobanked. After clinical assessment, a consensus diagnosis is made during a multidisciplinary diagnostic conference.
Diagnostic groups included in this study were adherent to the following clinical criteria: behavioral variant frontotemporal dementia (bvFTD) [3], dementia due to Alzheimer's disease (AD) [4], dementia with Lewy bodies (DLB) [5], logopenic variant primary progressive aphasia (lvPPA), non-fluent variant PPA (nfvPPA), and semantic variant PPA (svPPA) [6]. Although an international consensus has not yet been established, cases were classified as rtvFTD when they fulfilled the clinical and radiological criteria proposed by Ulugut et al., 2020 [7]. Patients were diagnosed with PPA 'not otherwise specified' (NOS) when they did not fully meet established any of the clinical criteria of Gorno-Tempini et al., 2011, but did exhibit a phenotype within the PPA spectrum as per original criteria by Mesulam et al., 2001. Patients were excluded from the AD and lvPPA groups if they exhibited negative AD biomarkers, as determined through either cerebrospinal fluid (CSF) analysis or amyloid PET imaging. Controls were selected from the ADC cohort among individuals diagnosed with subjective cognitive decline (SCD). To ensure an AD negative control group, SCD cases with evidence of positive AD biomarkers, defined as either abnormal CSF profiles or amyloid positivity on PET imaging, were excluded from the control sample. For all other diagnostic groups, AD biomarker status could be positive, negative, or unavailable (A+/A-/Missing): rtvFTD (9/36/6), lvPPA (35/0/3), nfvPPA (6/21/3), svPPA (12/43/11), PPA NOS (9/7/1), controls (0/733/236), bvFTD (41/154/35), AD (1543/0/207), DLB (120/86/50).

All participants included in the study presented here provided informed consent for the use of their clinical, biological and genetic data in research. The ADC cohort study is approved by its local medical ethics committee of the VU Medical Centre (file number 2016.061) and is performed in accordance with the ethical standards as laid down in the 1964 Declaration of Helsinki.

Participants from 23andMe

Summary statistics of Doust et al., 2022 [9], were provided by 23andMe Research Institute, a nonprofit medical research organization. All 23andMe participants provided online, opt-in informed consent to participate in research initiatives. 23andMe's protocol is approved by the external AAHRPP-accredited IRB, Ethical and Independent Review Services (www.eandireview.com).

**S2. Supplementary method on calculated polygenic risk scores for dyslexia**

LDpred2 PRS were computed using the R package bigsnpr (version 1.4.7), with the approach of a Genome Wide bed file and the infinitesimal model. LDpred2 uses a Bayesian approach to calculate a posterior mean effect size for each single nucleotide polymorphism based on prior GWAS effect sizes followed by shrinkage using Linkage Disequilibrium (LD) information [10]. We also used a set of HapMap3 SNPs (including 1,054,330 variants) in order to get adequate coverage of the genome, as recommended by the LDpred2 authors [10].

Since GWAS summary statistics were in build GRCh37 (hg19), we used R package liftOver and University of California (UCSC) chain file to transform coordinates of the summary statistics from hg19 to genotyped coordinates of hg38 (Alan Murphy, 2021; Bioconductor Package Maintainer, 2021). LDpred2 PRSs were computed using the R package bigsnpr (version 1.4.7), with the approach of a Genome Wide bed file and the infinitesimal model. In short, LDpred2 uses a Bayesian approach to calculate a posterior mean effect size for each SNP based on prior GWAS effect sizes followed by shrinkage using Linkage Disequilibrium (LD) information (Privé et al., 2020). Also, we used a set of HapMap3 SNPs (including 1,054,330 variants) in order to get adequate coverage of the genome, as recommended by the LDpred2 authors [10].

**Table S1.** Output linear regression analysis without stratification for sex, adjusted for age, sex and education.

| Reference Group | Diagnostic Group | Estimate | Standard Error | p-value |
|---|---|---|---|---|
| svPPA | rtvFTD | -0.253 | 0.098 | 9.45e-03* |
| | lvPPA | -0.179 | 0.106 | 9.19e-02 |
| | nfvPPA | -0.234 | 0.117 | 4.57e-02* |
| | PPA NOS | -0.477 | 0.141 | 7.41e-04* |
| | controls | -0.273 | 0.067 | 4.61e-05* |
| | bvFTD | -0.176 | 0.073 | 1.62e-02* |
| | AD | -0.218 | 0.066 | 9.59e-04* |
| | DLB | -0.229 | 0.073 | 1.64e-03* |
| nfvPPA | svPPA | 0.234 | 0.117 | 4.57e-02 |
| | rtvFTD | -0.019 | 0.122 | 8.76e-01 |
| | lvPPA | 0.056 | 0.129 | 6.66e-01 |
| | PPA NOS | -0.242 | 0.159 | 1.28e-01 |
| | controls | -0.039 | 0.1 | 6.95e-01 |
| | bvFTD | 0.058 | 0.104 | 5.76e-01 |
| | AD | 0.017 | 0.099 | 8.66e-01 |
| | DLB | 0.005 | 0.103 | 9.61e-01 |
| lvPPA | nfvPPA | -0.056 | 0.129 | 6.66e-01 |
| | svPPA | 0.179 | 0.106 | 9.19e-02 |
| | rtvFTD | -0.075 | 0.111 | 5.04e-01 |
| | PPA NOS | -0.298 | 0.151 | 4.84e-02 |
| | controls | -0.095 | 0.086 | 2.72e-01 |
| | bvFTD | 0.002 | 0.091 | 9.78e-01 |
| | AD | -0.039 | 0.085 | 6.46e-01 |
| | DLB | -0.05 | 0.09 | 5.76e-01 |
| rtvFTD | lvPPA | 0.075 | 0.111 | 5.04e-01 |
| | nfvPPA | 0.019 | 0.122 | 8.76e-01 |
| | svPPA | 0.253 | 0.098 | 9.45e-03 |
| | PPA NOS | -0.223 | 0.145 | 1.24e-01 |
| | controls | -0.02 | 0.075 | 7.89e-01 |
| | bvFTD | 0.077 | 0.081 | 3.41e-01 |
| | AD | 0.036 | 0.074 | 6.32e-01 |
| | DLB | 0.024 | 0.08 | 7.64e-01 |

Adjusted for age and education (years). *statistically significant.
Abbreviations: AD, Alzheimer’s disease; bvFTD, behavioral variant frontotemporal dementia; DLB, dementia with Lewy bodies; F, female; nfvPPA, non-fluent variant primary progressive aphasia; svPPA, semantic variant primary progressive aphasia; lvPPA, logopenic variant primary progressive aphasia; M, male; PPA NOS, primary progressive aphasia not otherwise specified;

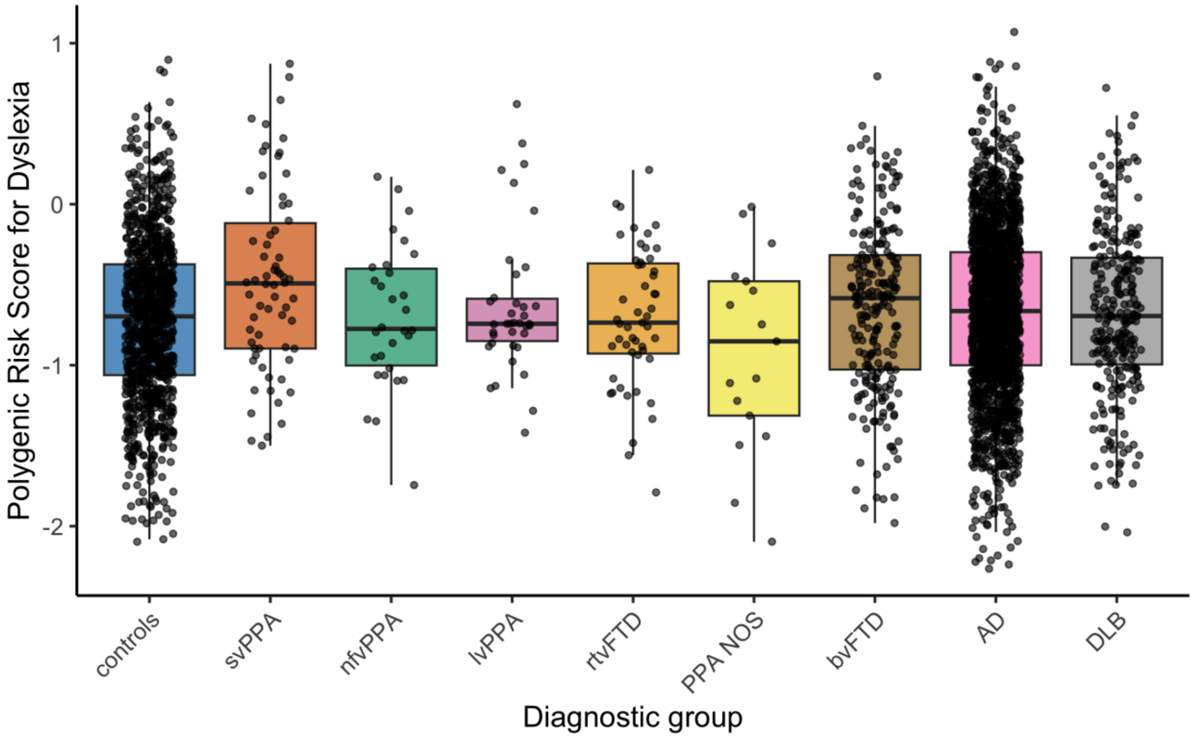


**Figure S3.** Polygenic risk scores for dyslexia per diagnostic group.
Polygenic risk scores for dyslexia is significantly higher in svPPA compared to all other groups (except lvPPA). No other significant differences were found.
Abbreviations: AD, Alzheimer's disease; bvFTD, behavioral variant frontotemporal dementia; DLB, dementia with Lewy bodies; nfvPPA, non-fluent variant primary progressive aphasia; svPPA, semantic variant primary progressive aphasia; lvPPA, logopenic variant primary progressive aphasia; PPA NOS, primary progressive aphasia not otherwise specified.

**Supplementary Material S2 – Sex specific PRS for dyslexia**

**Table S2.** Output linear regression analysis using sex-specific PRS for dyslexia within the corresponding sex group.

| Sex | Reference Group | Diagnostic Group | Estimate | Standard Error | p-value |
|---|---|---|---|---|---|
| M | svPPA | rtvFTD | -0.222 | 0.094 | 1.82e-02* |
| | | lvPPA | -0.253 | 0.11 | 2.09e-02* |
| | | nfvPPA | -0.13 | 0.132 | 3.25e-01 |
| | | controls | -0.22 | 0.067 | 1.08e-03* |
| | | PPA NOS | -0.348 | 0.152 | 2.23e-02* |
| | | bvFTD | -0.183 | 0.074 | 1.39e-02* |
| | | AD | -0.206 | 0.067 | 2.01e-03* |
| | | DLB | -0.258 | 0.071 | 3.09e-04* |
| F | svPPA | rtvFTD | -0.031 | 0.157 | 8.45e-01 |
| | | lvPPA | 0.034 | 0.14 | 8.08e-01 |
| | | nfvPPA | 0.14 | 0.151 | 3.54e-01 |
| | | controls | -0.02 | 0.096 | 8.33e-01 |
| | | PPA NOS | -0.158 | 0.186 | 3.98e-01 |
| | | bvFTD | 0.142 | 0.105 | 1.76e-01 |
| | | AD | 0.092 | 0.094 | 3.31e-01 |
| | | DLB | 0.098 | 0.117 | 4.04e-01 |
| M | nfvPPA | rtvFTD | -0.093 | 0.133 | 4.85e-01 |
| | | lvPPA | -0.124 | 0.144 | 3.91e-01 |
| | | controls | -0.091 | 0.116 | 4.33e-01 |
| | | PPA NOS | -0.219 | 0.179 | 2.22e-01 |
| | | bvFTD | -0.053 | 0.12 | 6.57e-01 |
| | | AD | -0.077 | 0.115 | 5.05e-01 |
| | | DLB | -0.129 | 0.118 | 2.75e-01 |
| F | nfvPPA | rtvFTD | -0.171 | 0.174 | 3.27e-01 |
| | | lvPPA | -0.106 | 0.158 | 5.04e-01 |
| | | controls | -0.16 | 0.122 | 1.89e-01 |
| | | PPA NOS | -0.298 | 0.2 | 1.37e-01 |
| | | bvFTD | 0.003 | 0.128 | 9.83e-01 |
| | | AD | -0.048 | 0.119 | 6.88e-01 |
| | | DLB | -0.042 | 0.137 | 7.60e-01 |
| M | lvPPA | rtvFTD | 0.031 | 0.111 | 7.80e-01 |
| | | controls | 0.033 | 0.09 | 7.14e-01 |
| | | PPA NOS | -0.095 | 0.163 | 5.61e-01 |
| | | bvFTD | 0.071 | 0.095 | 4.58e-01 |
| | | AD | 0.047 | 0.089 | 5.98e-01 |
| | | DLB | -0.005 | 0.092 | 9.57e-01 |
| F | lvPPA | rtvFTD | -0.065 | 0.165 | 6.93e-01 |
| | | controls | -0.054 | 0.108 | 6.14e-01 |
| | | PPA NOS | -0.192 | 0.192 | 3.19e-01 |
| | | bvFTD | 0.108 | 0.116 | 3.50e-01 |

| | | | | | |
|---|---|---|---|---|---|
| | | AD | 0.058 | 0.106 | 5.86e-01 |
| | | DLB | 0.063 | 0.126 | 6.15e-01 |
| M | rtvFTD | controls | 0.002 | 0.07 | 9.77e-01 |
| | | PPA NOS | -0.126 | 0.153 | 4.12e-01 |
| | | bvFTD | 0.04 | 0.077 | 6.05e-01 |
| | | AD | 0.016 | 0.069 | 8.18e-01 |
| | | DLB | -0.036 | 0.074 | 6.26e-01 |
| F | rtvFTD | controls | 0.011 | 0.129 | 9.34e-01 |
| | | PPA NOS | -0.127 | 0.205 | 5.37e-01 |
| | | bvFTD | 0.173 | 0.136 | 2.02e-01 |
| | | AD | 0.123 | 0.128 | 3.37e-01 |
| | | DLB | 0.129 | 0.146 | 3.78e-01 |

Adjusted for age and education (years). * statistically significant.
Abbreviations: AD, Alzheimer's disease; bvFTD, behavioral variant frontotemporal dementia; DLB, dementia with Lewy bodies; F, female; nfvPPA, non-fluent variant primary progressive aphasia; svPPA, semantic variant primary progressive aphasia; lvPPA, logopenic variant primary progressive aphasia; M, male; PPA NOS, primary progressive aphasia not otherwise specified

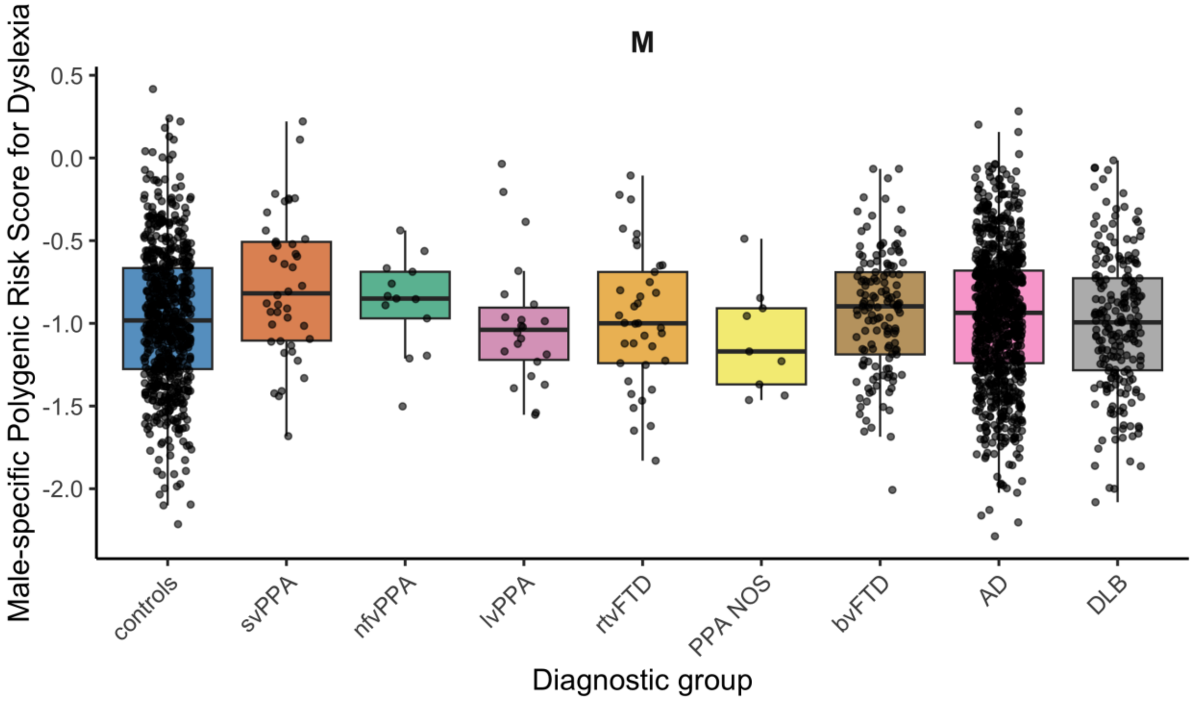


**Figure S1.** Male specific PRS for dyslexia per diagnostic group within male group.
Male specific PRS for dyslexia is significantly higher in the svPPA compared to all other groups except nfvPPA.
Abbreviations: AD, Alzheimer's disease; bvFTD, behavioral variant frontotemporal dementia; DLB, dementia with Lewy bodies; nfvPPA, non-fluent variant primary progressive aphasia; svPPA, semantic variant primary progressive aphasia; lvPPA, logopenic variant primary progressive aphasia; PPA NOS, primary progressive aphasia not otherwise specified.

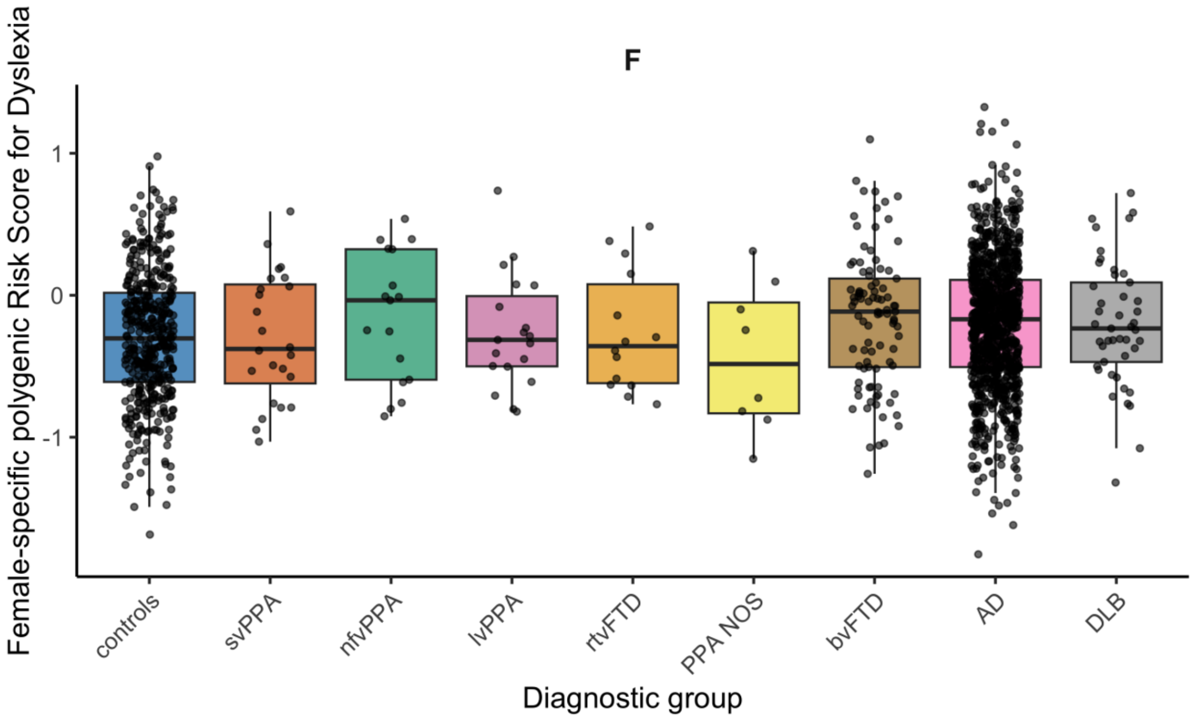


**Figure S2.** Female specific PRS for dyslexia per diagnostic group within female group.
No significant differences were found for polygenic risk scores for dyslexia between diagnostic groups.
Abbreviations: AD, Alzheimer's disease; bvFTD, behavioral variant frontotemporal dementia; DLB, dementia with Lewy bodies; nfvPPA, non-fluent variant primary progressive aphasia; svPPA, semantic variant primary progressive aphasia; lvPPA, logopenic variant primary progressive aphasia; PPA NOS, primary progressive aphasia not otherwise specified.

**Supplementary Material S3**

Linear regression models were run without adjustment for education (in years).

**Table S3**. Linear Regression analysis stratified for sex, adjusted for age.

| Sex | Reference Group | Diagnostic Group | Estimate | Standard Error | p-value |
|---|---|---|---|---|---|
| M | svPPA | rtvFTD | -0.369 | 0.116 | 1.54e-03* |
| | | lvPPA | -0.257 | 0.143 | 7.22e-02 |
| | | nfvPPA | -0.256 | 0.164 | 1.18e-01 |
| | | PPA NOS | -0.515 | 0.189 | 6.63e-03* |
| | | controls | -0.329 | 0.083 | 6.92e-05* |
| | | bvFTD | -0.306 | 0.091 | 7.76e-04* |
| | | AD | -0.337 | 0.082 | 3.80e-05* |
| | | DLB | -0.346 | 0.087 | 7.77e-05* |
| F | svPPA | rtvFTD | 0.017 | 0.174 | 9.23e-01 |
| | | lvPPA | 0.032 | 0.159 | 8.41e-01 |
| | | nfvPPA | -0.085 | 0.164 | 6.04e-01 |
| | | PPA NOS | -0.303 | 0.211 | 1.51e-01 |
| | | controls | -0.077 | 0.109 | 4.82e-01 |
| | | bvFTD | 0.067 | 0.118 | 5.71e-01 |
| | | AD | 0.037 | 0.107 | 7.30e-01 |
| | | DLB | 0.092 | 0.131 | 4.83e-01 |
| M | nfvPPA | rtvFTD | -0.113 | 0.166 | 4.98e-01 |
| | | lvPPA | -0.001 | 0.186 | 9.96e-01 |
| | | PPA NOS | -0.259 | 0.224 | 2.47e-01 |
| | | controls | -0.073 | 0.145 | 6.14e-01 |
| | | bvFTD | -0.05 | 0.15 | 7.38e-01 |
| | | AD | -0.081 | 0.144 | 5.75e-01 |
| | | DLB | -0.09 | 0.147 | 5.42e-01 |
| F | nfvPPA | rtvFTD | 0.102 | 0.187 | 5.87e-01 |
| | | lvPPA | 0.117 | 0.173 | 4.98e-01 |
| | | PPA NOS | -0.218 | 0.222 | 3.25e-01 |
| | | controls | 0.008 | 0.13 | 9.49e-01 |
| | | bvFTD | 0.152 | 0.137 | 2.67e-01 |
| | | AD | 0.122 | 0.127 | 3.36e-01 |
| | | DLB | 0.177 | 0.147 | 2.29e-01 |
| M | lvPPA | rtvFTD | -0.112 | 0.146 | 4.43e-01 |
| | | PPA NOS | -0.258 | 0.209 | 2.16e-01 |
| | | controls | -0.072 | 0.121 | 5.51e-01 |
| | | bvFTD | -0.049 | 0.127 | 6.97e-01 |
| | | AD | -0.08 | 0.12 | 5.04e-01 |
| | | DLB | -0.089 | 0.123 | 4.70e-01 |
| F | lvPPA | rtvFTD | -0.015 | 0.183 | 9.34e-01 |
| | | PPA NOS | -0.335 | 0.218 | 1.24e-01 |
| | | controls | -0.109 | 0.123 | 3.75e-01 |
| | | bvFTD | 0.035 | 0.131 | 7.88e-01 |
| | | AD | 0.005 | 0.12 | 9.68e-01 |
| | | DLB | 0.06 | 0.141 | 6.72e-01 |

| | | | | | |
|---|---|---|---|---|---|
| M | rtvFTD | PPA NOS | -0.146 | 0.192 | 4.45e-01 |
| | | controls | 0.04 | 0.088 | 6.52e-01 |
| | | bvFTD | 0.063 | 0.096 | 5.13e-01 |
| | | AD | 0.032 | 0.087 | 7.14e-01 |
| | | DLB | 0.023 | 0.092 | 8.06e-01 |
| F | rtvFTD | PPA NOS | -0.32 | 0.23 | 1.63e-01 |
| | | controls | -0.094 | 0.141 | 5.06e-01 |
| | | bvFTD | 0.05 | 0.148 | 7.34e-01 |
| | | AD | 0.02 | 0.14 | 8.86e-01 |
| | | DLB | 0.075 | 0.159 | 6.37e-01 |

*statistically significant.

Abbreviations: AD, Alzheimer's disease; bvFTD, behavioral variant frontotemporal dementia; DLB, dementia with Lewy bodies; F, female; nfvPPA, non-fluent variant primary progressive aphasia; svPPA, semantic variant primary progressive aphasia; lvPPA, logopenic variant primary progressive aphasia; M, male; PPA NOS, primary progressive aphasia not otherwise specified.

**Table S4**. Linear regression analysis without sex stratification, adjusted for age.

| Reference Group | Diagnostic Group | Estimate | Standard Error | p-value |
|---|---|---|---|---|
| svPPA | rtvFTD | -0.231 | 0.096 | 1.66e-02* |
| | lvPPA | -0.157 | 0.105 | 1.36e-01 |
| | nfvPPA | -0.23 | 0.114 | 4.40e-02* |
| | PPA NOS | -0.45 | 0.141 | 1.38e-03* |
| | controls | -0.239 | 0.066 | 2.99e-04* |
| | bvFTD | -0.167 | 0.072 | 2.04e-02* |
| | AD | -0.194 | 0.065 | 2.82e-03* |
| | DLB | -0.204 | 0.072 | 4.31e-03* |
| nfvPPA | svPPA | 0.23 | 0.114 | 4.40e-02 |
| | rtvFTD | -0.001 | 0.119 | 9.90e-01 |
| | lvPPA | 0.072 | 0.126 | 5.66e-01 |
| | PPA NOS | -0.221 | 0.157 | 1.60e-01 |
| | controls | -0.009 | 0.096 | 9.24e-01 |
| | bvFTD | 0.062 | 0.101 | 5.37e-01 |
| | AD | 0.036 | 0.095 | 7.08e-01 |
| | DLB | 0.025 | 0.1 | 8.02e-01 |
| lvPPA | nfvPPA | -0.072 | 0.126 | 5.66e-01 |
| | svPPA | 0.157 | 0.105 | 1.36e-01 |
| | rtvFTD | -0.074 | 0.111 | 5.06e-01 |
| | PPA NOS | -0.293 | 0.151 | 5.20e-02 |
| | controls | -0.082 | 0.086 | 3.43e-01 |
| | bvFTD | -0.01 | 0.091 | 9.09e-01 |
| | AD | -0.037 | 0.085 | 6.64e-01 |
| | DLB | -0.047 | 0.09 | 5.99e-01 |
| rtvFTD | lvPPA | 0.074 | 0.111 | 5.06e-01 |
| | nfvPPA | 0.001 | 0.119 | 9.90e-01 |
| | svPPA | 0.231 | 0.096 | 1.66e-02 |
| | PPA NOS | -0.219 | 0.145 | 1.30e-01 |
| | controls | -0.008 | 0.074 | 9.17e-01 |
| | bvFTD | 0.064 | 0.08 | 4.27e-01 |
| | AD | 0.037 | 0.074 | 6.15e-01 |
| | DLB | 0.027 | 0.079 | 7.39e-01 |

*statistically significant.

Abbreviations: AD, Alzheimer's disease; bvFTD, behavioral variant frontotemporal dementia; DLB, dementia with Lewy bodies; F, female; nfvPPA, non-fluent variant primary progressive aphasia; svPPA, semantic variant primary progressive aphasia; lvPPA, logopenic variant primary progressive aphasia; M, male; PPA NOS, primary progressive aphasia not otherwise specified.
